# Population-based Ethnoracial Disparity in Life’s Essential 8 Vascular Health, Cerebral Small Vessel Disease & Cognition

**DOI:** 10.64898/2026.10.09.26365181

**Authors:** Nikita K. Husein, Mai P. Ho, Keshuo Lin, David K. E. Chan, Shizuka Hayashi, Sonal P. Sachdev, Tahereh Hassanzadeh, Jiyang Jiang, Perminder S. Sachdev, Wei Wen

## Abstract

**Background:** Whether ethnoracial differences in cerebral small vessel disease (cSVD) appear in multimarker burden, particular MRI manifestations, or both, and how they relate to Life’s Essential 8 (LE8) cardiovascular health and cognition, is unclear.

**Methods:** This cross-sectional study included 44,982 participants and an age- and sex-matched subsample of 1,426 with reviewed lacune and cerebral microbleed (CMB) ratings. White, South Asian, Black, Chinese and Mixed participants were compared on 0–2 (white matter hyperintensity [WMH]/enlarged perivascular space [ePVS]) and 0–4 (adding CMB and lacunes) composite scores and each marker. LE8 was related to WMH and composite burden, and cognitive means were compared by group. Models were age-, sex- and center-adjusted with false-discovery-rate correction.

**Results:** Both composite scores differed across groups (0–2: P=0.00010; 0–4: P=0.0011), with greater burden in Black than South Asian (0–4: OR 1.66; 95% CI, 1.21–2.28) and Chinese participants (OR 2.36; 1.51–3.70), whereas Black–White contrasts were not detected. Black participants had higher WMH volume than White participants (β=0.209; 0.110–0.308), robust to deprivation adjustment and segmentation method; South Asian participants had fewer ePVS (0.82; 0.72–0.93). No ethnoracial-group differences in lacune or CMB counts were detected after correction. Higher LE8 was associated with lower WMH volume (β=−0.13 per SD; −0.17 to −0.09) and lower two-marker burden (OR 0.89 per SD; 0.87–0.91) without detected ethnoracial moderation. Black participants, who had the least favorable LE8 scores, and South Asian participants had lower global cognition than White participants (z −0.76 and −0.57 versus 0.24) despite differing WMH profiles; the pooled WMH–cognition association was null.

**Conclusions:** Ethnoracial differences in multimarker cSVD burden were evident but marker-dependent: WMH showed the clearest Black–White difference; focal-lesion contrasts were inconclusive. Better cardiovascular health related to lower WMH and aggregate cSVD burden across groups, whereas cognitive differences did not track cSVD burden, supporting prospective prevention and cognition studies.

## Introduction

Cerebral small vessel disease (cSVD) is a major contributor to ischemic and hemorrhagic stroke and vascular cognitive impairment. Its MRI markers—white matter hyperintensities (WMH), lacunes, cerebral microbleeds (CMB), and enlarged perivascular spaces (ePVS)—represent related but distinct manifestations of small-vessel injury and are not clinically interchangeable. Even without clinical symptoms, greater MRI cSVD burden is associated with future stroke and cognitive outcomes.^1–5^ UK stroke-register studies have documented differences in stroke incidence across self-reported ethnic groups.^6,7^ Determining whether subclinical vascular brain injury also differs across groups may therefore inform cerebrovascular epidemiology and prevention research. Such comparisons, however, must distinguish observed imaging differences from the vascular, socioeconomic, life-course, selection and measurement processes that can shape them.

Available evidence suggests that ethnoracial differences in cSVD are heterogeneous across markers and populations rather than forming a consistent burden hierarchy. US studies have reported greater WMH burden in Black adults, with variable attenuation after socioeconomic and cardiovascular adjustment.^8–10^ A 2026 systematic review and meta-analysis found ethnic variation across WMH, lacunes, CMBs and vascular-risk profiles, but ePVS could not be meta-analyzed because ethnically diverse ePVS data were insufficient.^11^ A broader global synthesis likewise shows substantial heterogeneity in cSVD burden.^12^ An earlier UK Biobank analysis combined several ethnic groups in a binary comparison, limiting interpretation for named groups.^13^ In SABRE, South Asian and European participants had similar WMH distributions but different risk-factor associations; Chinese–White comparisons and the Northern Manhattan Study further show that findings for WMH cannot simply be extended to other cSVD markers.^14–16^

We therefore asked whether ethnoracial differences in cSVD in UK Biobank appear as differences in overall multimarker burden, as differences in particular MRI manifestations, or both, and whether cardiovascular-health associations differ across groups. Composite total-SVD scores are an established pragmatic summary of aggregate burden,^17^ whereas STRIVE-2 and recent multimarker work, including in Stroke, support evaluating the individual manifestations jointly because they represent related but non-identical forms of small-vessel injury.^3,18^ Our 2026 systematic review indicates that ethnoracial differences are heterogeneous across markers rather than forming one universal hierarchy.^11^ We compared White, South Asian, Black, Chinese and Mixed participants on two study-specific composite scores and on their constituent markers, treating self-reported identities as markers of socially patterned vascular and life-course exposures rather than inherited susceptibility.^19,20^ This framing follows the National Institute on Minority Health and Health Disparities Research Framework, which locates group differences in biological, behavioral, physical-environmental, sociocultural and healthcare domains across levels of influence.^21^ Socioeconomic adjustment, cardiovascular-health associations and an imaging-selection analysis provide the measured exposure and sampling context.

## Methods

### Study design and ethics

This cross-sectional imaging study follows STROBE recommendations.^22^ UK Biobank recruited adults aged 40–69 years in 2006–2010; 3-T MRI was acquired from 2014 onward. UK Biobank has approval from the North West Multi-center Research Ethics Committee as a Research Tissue Bank. All participants provided written informed consent at recruitment and at the neuroimaging assessment. This analysis was conducted under approved UK Biobank application 98013 and did not require separate ethical approval or additional participant consent.

### Data Availability

Individual participant data cannot be redistributed directly by the authors under the terms of UK Biobank access. Bona fide researchers may apply for the same data through the UK Biobank Access Management System (https://www.ukbiobank.ac.uk). Derived analysis code and nonidentifiable, aggregate summary outputs may be made available on reasonable request to the corresponding author.

### Participants

The full sample comprised 44,982 participants, with 44,580 assigned to an ethnic group: White 43,550, South Asian 393, Black 288, Chinese 131 and Mixed 218. The matched sample (N=1,426) comprised White 396, South Asian 393, Black 290, Chinese 131 and Mixed 216 participants. Its White comparison group was age- and sex-frequency-matched to the pooled distribution of South Asian, Black, Chinese and Mixed participants, not to each group separately. Imaging center and deprivation were not frequency-matching variables. The White groups had mean ages of 64.9 years in the full sample and 61.7 years in the matched sample (Table 1).

**Table 1.**
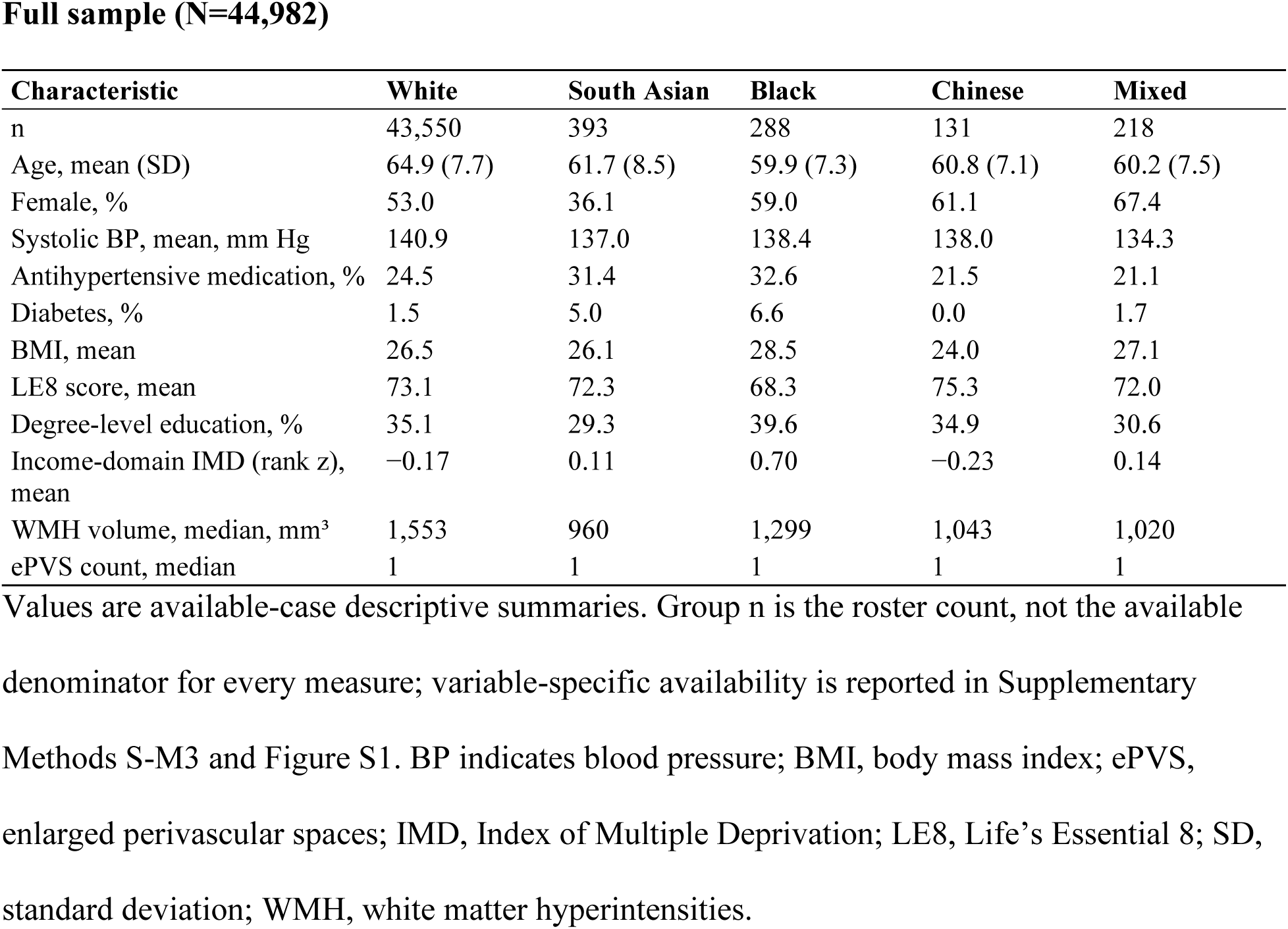
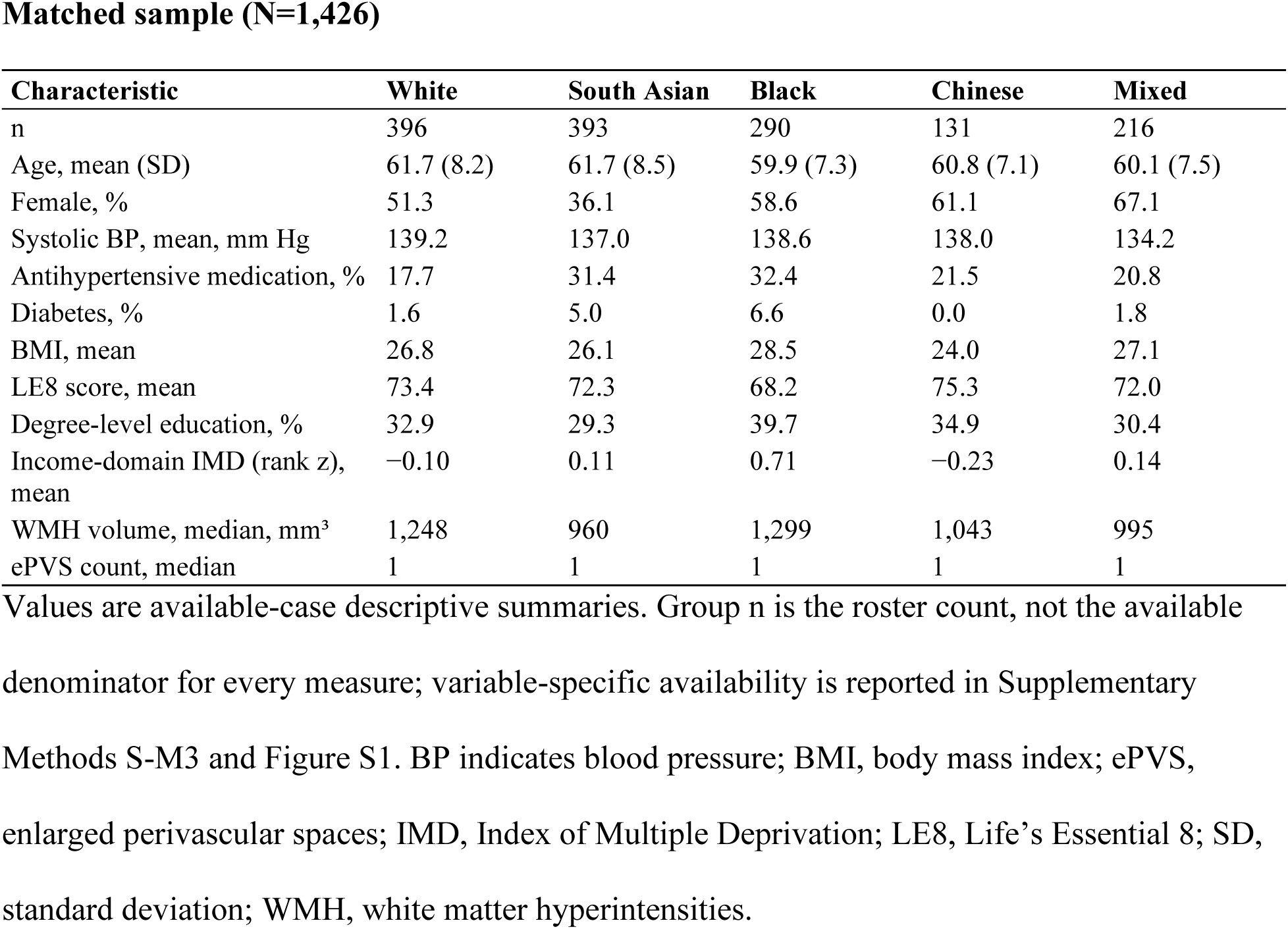
Participant characteristics.

**Full sample (N=44,982)**
| Characteristic | White | South Asian | Black | Chinese | Mixed |
| --- | --- | --- | --- | --- | --- |
| n | 43,550 | 393 | 288 | 131 | 218 |
| Age, mean (SD) | 64.9 (7.7) | 61.7 (8.5) | 59.9 (7.3) | 60.8 (7.1) | 60.2 (7.5) |
| Female, % | 53.0 | 36.1 | 59.0 | 61.1 | 67.4 |
| Systolic BP, mean, mm Hg | 140.9 | 137.0 | 138.4 | 138.0 | 134.3 |
| Antihypertensive medication, % | 24.5 | 31.4 | 32.6 | 21.5 | 21.1 |
| Diabetes, % | 1.5 | 5.0 | 6.6 | 0.0 | 1.7 |
| BMI, mean | 26.5 | 26.1 | 28.5 | 24.0 | 27.1 |
| LE8 score, mean | 73.1 | 72.3 | 68.3 | 75.3 | 72.0 |
| Degree-level education, % | 35.1 | 29.3 | 39.6 | 34.9 | 30.6 |
| Income-domain IMD (rank z), mean | −0.17 | 0.11 | 0.70 | −0.23 | 0.14 |
| WMH volume, median, mm<sup>3</sup> | 1,553 | 960 | 1,299 | 1,043 | 1,020 |
| ePVS count, median | 1 | 1 | 1 | 1 | 1 |

**Matched sample (N=1,426)**
| Characteristic | White | South Asian | Black | Chinese | Mixed |
| --- | --- | --- | --- | --- | --- |
| n | 396 | 393 | 290 | 131 | 216 |
| Age, mean (SD) | 61.7 (8.2) | 61.7 (8.5) | 59.9 (7.3) | 60.8 (7.1) | 60.1 (7.5) |
| Female, % | 51.3 | 36.1 | 58.6 | 61.1 | 67.1 |
| Systolic BP, mean, mm Hg | 139.2 | 137.0 | 138.6 | 138.0 | 134.2 |
| Antihypertensive medication, % | 17.7 | 31.4 | 32.4 | 21.5 | 20.8 |
| Diabetes, % | 1.6 | 5.0 | 6.6 | 0.0 | 1.8 |
| BMI, mean | 26.8 | 26.1 | 28.5 | 24.0 | 27.1 |
| LE8 score, mean | 73.4 | 72.3 | 68.2 | 75.3 | 72.0 |
| Degree-level education, % | 32.9 | 29.3 | 39.7 | 34.9 | 30.4 |
| Income-domain IMD (rank z), mean | -0.10 | 0.11 | 0.71 | -0.23 | 0.14 |
| WMH volume, median, mm<sup>3</sup> | 1,248 | 960 | 1,299 | 1,043 | 995 |
| ePVS count, median | 1 | 1 | 1 | 1 | 1 |

Adjusted matched-sample group profiles are presented in Table 2. These are overlapping analysis levels: South Asian and Chinese participants are identical across levels, while Black and Mixed membership differs only through the roster decisions described below. Automated-marker analyses at the matched-sample level therefore assess sensitivity to the White comparison group and cohort implementation, not independent confirmation.

**Table 2.** Adjusted clinical profiles in the matched sample (N=1,426)

| Measure | Model n | White | South Asian | Black | Chinese | Mixed |
| --- | --- | --- | --- | --- | --- | --- |
| **Cardiovascular health** |  |  |  |  |  |  |
| LE8 total score (0–100) | 1423 | 73.5 (72.3, 74.7) | 73.0 (71.8, 74.2) | 67.7 (66.3, 69.1) | 74.9 (72.8, 77.0) | 71.2 (69.6, 72.8) |
| Systolic blood pressure (mm Hg) | 1410 | 138.7 (136.9, 140.5) | 135.8 (134.0, 137.7) | 139.8 (137.7, 142.0) | 138.7 (135.5, 141.9) | 135.6 (133.2, 138.1) |
| Body mass index (kg/m<sup>2</sup>) | 1424 | 26.8 (26.4, 27.3) | 26.2 (25.7, 26.6) | 28.4 (27.9, 28.9) | 24.0 (23.2, 24.8) | 27.1 (26.5, 27.7) |
| **cSVD MRI** |  |  |  |  |  |  |
| UBO WMH volume, log(mm<sup>3</sup>+1) | 1411 | 7.17 (7.09, 7.25) | 7.01 (6.92, 7.09) | 7.28 (7.18, 7.38) | 7.07 (6.93, 7.21) | 7.06 (6.95, 7.17) |
| WMH lesion count at a given WMH volume | 1425 | 30.3 (28.9, 31.8) | 34.3 (32.6, 36.0) | 36.5 (34.5, 38.6) | 33.6 (31.0, 36.5) | 32.4 (30.4, 34.6) |
| ePVS count | 1423 | 1.58 (1.38, 1.81) | 1.38 (1.19, 1.59) | 1.94 (1.65, 2.27) | 1.42 (1.12, 1.81) | 1.54 (1.28, 1.86) |
| **Cognition** |  |  |  |  |  |  |
| Global cognition (z) | 1043 | 0.24 (0.13, 0.35) | −0.57 (−0.69, −0.45) | −0.76 (−0.90, −0.62) | −0.12 (−0.32, 0.08) | 0.01 (−0.15, 0.17) |
| Processing speed (z) | 1039 | 0.25 (0.15, 0.36) | −0.42 (−0.54, −0.31) | −0.51 (−0.64, −0.38) | 0.16 (−0.03, 0.35) | 0.05 (−0.09, 0.20) |
| Memory (z) | 1008 | 0.11 (−0.01, 0.24) | −0.27 (−0.40, −0.13) | −0.39 (−0.55, −0.23) | 0.27 (0.05, 0.50) | 0.02 (−0.16, 0.20) |
| Executive function (z) | 1017 | 0.18 (0.08, 0.28) | −0.69 (−0.80, −0.58) | −0.82 (−0.94, −0.69) | −0.65 (−0.83, −0.47) | −0.05 (−0.19, 0.09) |

Values are saved estimated marginal means (95% CI); n denotes model complete cases. Adjustment: cardiovascular measures and cognition, age and sex only; WMH volume, age, sex, center and ICV; WMH count, age, sex, center and log WMH volume; ePVS, age, sex and center. Count estimates are response-scale predictions. Abbreviations: CI, confidence interval; cSVD, cerebral small vessel disease; ePVS, enlarged perivascular spaces; ICV, intracranial volume; LE8, Life's Essential 8; UBO, UBO Detector (automated WMH segmentation pipeline); WMH, white matter hyperintensities.

The matched-sample analyses used the complete 1,426-participant roster to maximize statistical power and precision for group comparisons. White comparison participants were selected using seeded age-by-sex quotas without access to imaging outcomes. Lacune and CMB assessment followed a single common workflow across this sample. Sample composition and outcome-specific denominators are detailed in Supplementary Methods S-M1–S-M3 and Figure S1.

### Self-reported ethnic group

Ethnic group was recorded in UK Biobank Field 21000 at up to four assessments. The established algorithm used the first classifiable response, skipped nonresponses and did not assign nonspecific responses to a study group. The matched-sample roster retained two additional Black participants whose responses were unclassified by the algorithm and excluded two participants with absent or inconsistent Mixed classifications. These decisions explain the small differences in Black and Mixed group counts across the two levels. Ethnic group was the exposure, not an adjustment covariate. The five categories describe social identities and conceal substantial within-group heterogeneity; they are not measures of inherited susceptibility.^19,20^ Reporting of ethnic group follows the AMA Manual of Style guidance on race and ethnicity.^23^ Groups are listed with the White comparison group first and then in the cohort’s established order.

### MRI markers and focal-lesion ascertainment

WMH volume was quantified with UBO Detector for the main analysis, using a cluster-based pipeline based on T1 and FLAIR images that also provides lesion counts; the standard UK Biobank Brain Intensity AbNormality Classification Algorithm (BIANCA) measure was used only in sensitivity analyses to check the UBO findings.^24,25^ UBO Detector was retained as the primary method because an independent comparison of three freely available pipelines against manual reference segmentations found that it performed comparably to BIANCA without requiring a training set and agreed strongly with Fazekas ratings, whereas FreeSurfer underestimated WMH volume and BIANCA produced outlying volumes.^26^ Volumes were analyzed as log(x+1). ePVS were segmented with Long-PVSUNet, an in-house deep-learning pipeline, from T1-weighted and T2-weighted MRI, yielding basal ganglia, centrum semiovale, and total counts.^27^ Total ePVS count was a core automated marker. Regional measures included periventricular and deep WMH, and basal ganglia and centrum semiovale ePVS. Peak width of skeletonized mean diffusivity (PSMD) and difference in distribution functions (DDF) were secondary diffusion measures of white matter integrity.^28,29^ Both are described further in Supplementary Methods S-M9. Structural brain volumes, cortical thickness and surface area were analyzed separately as broader brain measures, not classified as cSVD markers. Group-stratified validation against manual reference segmentations was not available. The alternative WMH method was not treated as a reference standard.

Table 3 summarizes focal-lesion assessment and validation. Lacunes were rated de novo by direct visual inspection using Standards for Reporting Vascular Changes on Neuroimaging (STRIVE-2) criteria, excluding the hippocampal region, with a lesion counted as present at a likelihood rating of 2 or above.^3^ All rated lacunes underwent two-rater review and radiologist validation under the same blinded workflow across the matched sample. Raters did not see participants’ ethnoracial-group information during rating. The archived reliability assessment included an independent second rating of 78 enriched scans (Cohen κ=0.72; raw agreement 94.9%) with radiologist validation.

**Table 3.**
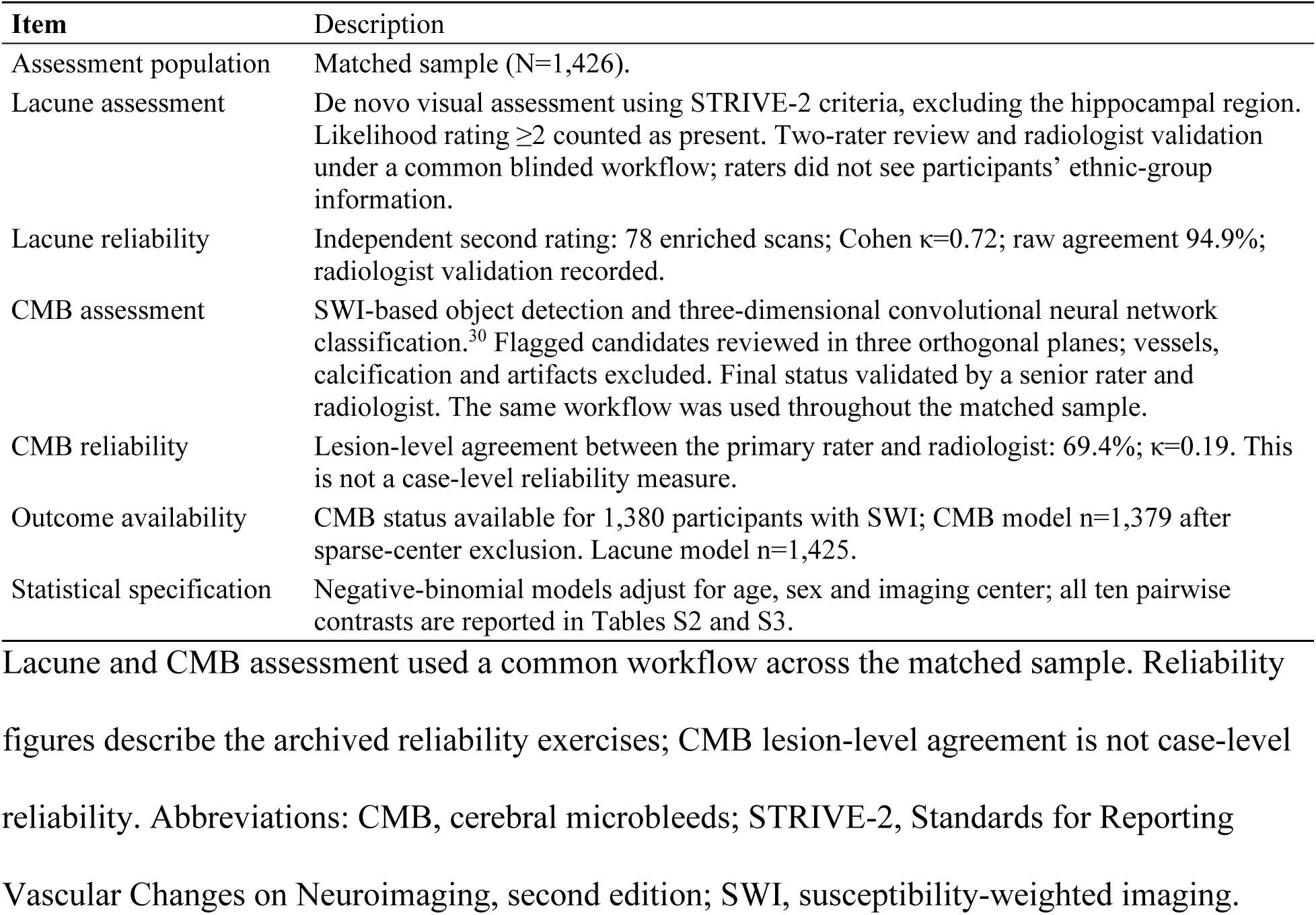
Focal-lesion assessment and validation.

CMB candidates were identified on susceptibility-weighted imaging (SWI) using an in-house deep-learning framework combining object detection with three-dimensional convolutional neural network classification.^30^ Each flagged candidate was reviewed in three orthogonal planes; vessels, calcification and artifacts were excluded, and final CMB status was validated by a senior rater and radiologist. This workflow was applied across the matched sample. The archived lesion-level agreement between the primary rater and radiologist was 69.4% (κ=0.19). CMB status was available for 1,380 participants with SWI, with 1,379 included after the sparse-center exclusion. Assessment procedures are summarized in Table 3 and Supplementary Methods S-M4.

### Cardiovascular, socioeconomic and cognitive measures

Cardiovascular health was summarized by the American Heart Association Life’s Essential 8 score (LE8), with higher scores indicating better health.^31^ Vascular covariates were systolic blood pressure, body mass index, diabetes and smoking. The income domain of the Index of Multiple Deprivation (IMD), harmonized within nation as a rank-based z score, was the primary socioeconomic adjustment; supplementary models included household income and highest qualification. Imaging selection was examined by comparing deprivation in imaged and eligible never-imaged participants. Global cognition and processing-speed, memory and executive measures were secondary descriptive outcomes.^32,33^ The cognitive adjustment hierarchy is reported in the supplement to assess measured educational and socioeconomic context, not to explain the principal imaging contrasts.

### Statistical analysis

Linear regression was used for log WMH volumes and diffusion measures; negative-binomial regression was used for counts. Models included age, sex and imaging center, with intracranial volume (ICV) for volume measures. WMH lesion-count models additionally included WMH volume: their incidence rate ratios (IRRs) represent lesion number at a given volume, not incident disease or a causal effect. The established sparse-center rule excluded centers contributing fewer than five participants to a model. Analyses used available complete cases for the relevant outcome and covariates, with outcome-specific denominators reported; missing CMB status was not coded as absence. Tests of signed regression coefficients and pairwise contrasts were 2-sided; omnibus tests used the model-specific reference distribution. No formal a priori power calculation was performed; the complete matched sample was used to maximize statistical power and precision for group comparisons.

The full sample is the main analysis set for core automated markers (Table 4; Figure 1); matched-sample estimates assess sensitivity to the White comparison group (Table S1). Lacunes, CMB and the four-marker composite use the matched sample, where focal-lesion assessments are available. The two-marker composite uses both analysis levels. This hierarchy follows marker availability and sampling design. For the main automated-marker comparisons and the matched-sample composite comparisons, Benjamini–Hochberg correction was applied across all ten pairs within each marker or score, analysis level and adjustment stage. The adjusted P value is reported as q; q<0.05 (false discovery rate [FDR]) defines detection within these families, alongside effect size and 95% CI. This is not study-wide error control. Other outcome, interaction and sensitivity families retain their separately defined corrections. Table 5 reports lacune and CMB contrasts under the same all-pairwise correction; these comparisons remain exploratory because focal-lesion events are sparse.

**Figure 1.**
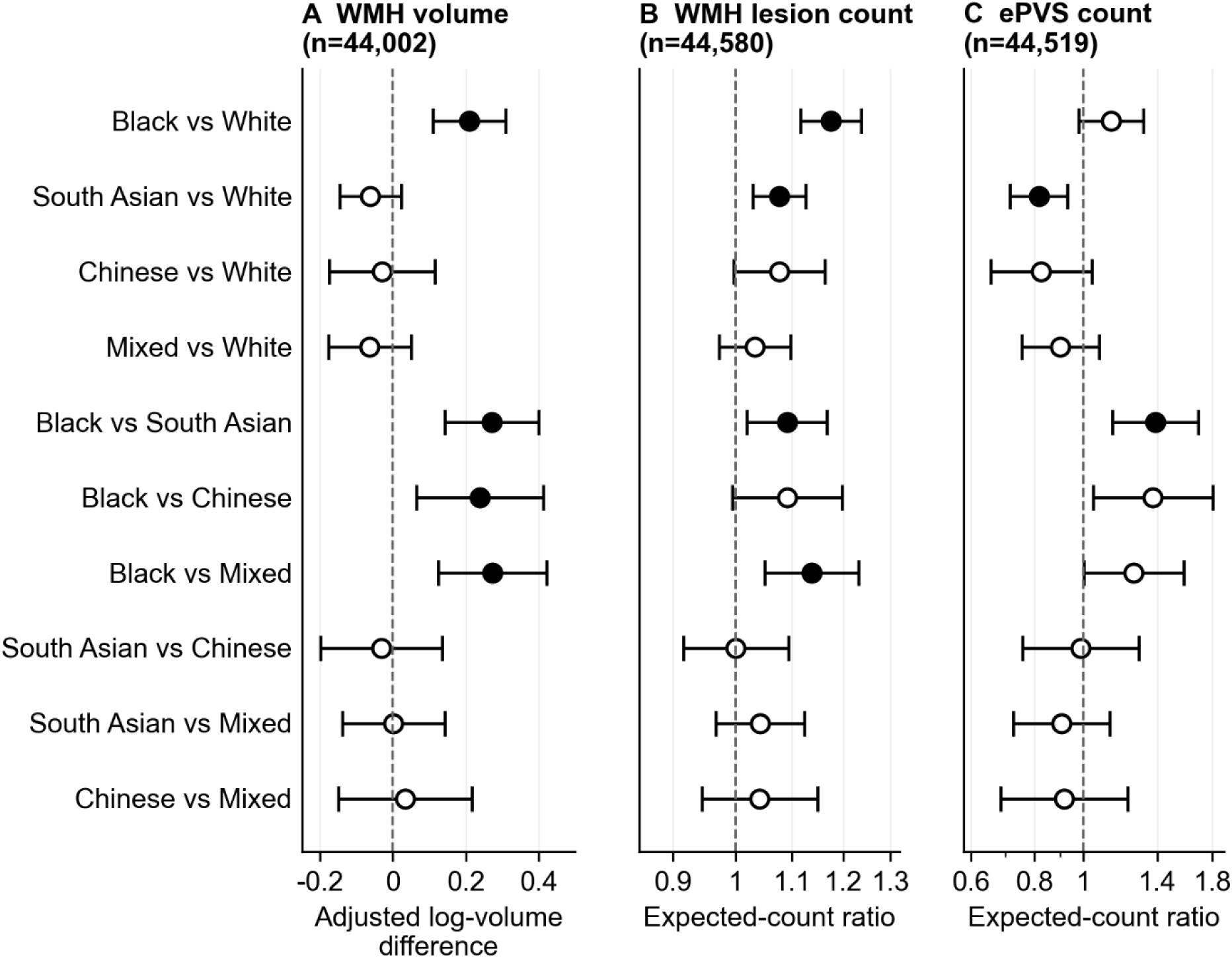
All ten pairwise contrasts for the core automated cSVD markers in the full sample. A, Adjusted differences in UBO Detector log(WMH+1) volume (model n=44,002). B and C, Incidence rate ratios (IRR) for WMH lesion count (model n=44,580) and ePVS count (model n=44,519). Models include age, sex and imaging center, with intracranial volume for WMH volume and WMH-volume adjustment for lesion count. Filled points indicate q<0.05 after Benjamini–Hochberg correction within each marker across all 10 pairwise contrasts; open points indicate q≥0.05. Horizontal lines show 95% CIs; the dashed line marks no difference. Contrasts compare the first named group with the second. cSVD indicates cerebral small vessel disease; ePVS, enlarged perivascular spaces; WMH, white matter hyperintensities.

**Table 4.** Automated cSVD markers in the full sample Main adjustment model.

| Group vs White | Model n | Estimate (95% CI) | P | q |
| --- | --- | --- | --- | --- |
| **WMH volume (UBO, log); $\beta$** |  |  |  |  |
| Black | 44,002 | 0.209 (0.110 to 0.308) | $3.5 \times 10^{-5}$ | 0.000195 |
| South Asian | 44,002 | -0.062 (-0.147 to 0.023) | 0.153 | 0.306 |
| Chinese | 44,002 | -0.030 (-0.174 to 0.115) | 0.688 | 0.796 |
| Mixed | 44,002 | -0.063 (-0.176 to 0.049) | 0.269 | 0.448 |
| **WMH volume (BIANCA, sensitivity); $\beta$** |  |  |  |  |
| Black | 44,002 | 0.311 (0.211 to 0.412) | $1.1 \times 10^{-9}$ | $1.1 \times 10^{-8}$ |
| South Asian | 44,002 | 0.074 (-0.012 to 0.159) | 0.0920 | 0.153 |
| Chinese | 44,002 | 0.144 (-0.002 to 0.290) | 0.0535 | 0.125 |
| Mixed | 44,002 | 0.054 (-0.059 to 0.168) | 0.350 | 0.437 |
| **WMH lesion count; IRR** |  |  |  |  |
| Black | 44,580 | 1.18 (1.12 to 1.24) | $8.19 \times 10^{-10}$ | $8.19 \times 10^{-9}$ |
| South Asian | 44,580 | 1.08 (1.03 to 1.13) | 0.00118 | 0.00454 |
| Chinese | 44,580 | 1.08 (1.00 to 1.16) | 0.0613 | 0.104 |
| Mixed | 44,580 | 1.03 (0.97 to 1.10) | 0.291 | 0.363 |
| **ePVS count; IRR** |  |  |  |  |
| Black | 44,519 | 1.13 (0.98 to 1.31) | 0.0914 | 0.173 |
| South Asian | 44,519 | 0.82 (0.72 to 0.93) | 0.00256 | 0.0128 |
| Chinese | 44,519 | 0.83 (0.66 to 1.04) | 0.104 | 0.173 |
| Mixed | 44,519 | 0.90 (0.76 to 1.07) | 0.247 | 0.353 |

**Additional income-domain IMD adjustment**
| Group vs White | Model n | Estimate (95% CI) | P | q |
| --- | --- | --- | --- | --- |
| **WMH volume (UBO, log); $\beta$** |  |  |  |  |
| Black | 42,962 | 0.178 (0.077 to 0.279) | 0.000552 | 0.00276 |
| South Asian | 42,962 | -0.062 (-0.148 to 0.025) | 0.162 | 0.323 |
| Chinese | 42,962 | -0.034 (-0.181 to 0.113) | 0.651 | 0.832 |
| Mixed | 42,962 | -0.073 (-0.187 to 0.041) | 0.208 | 0.347 |
| **WMH volume (BIANCA, sensitivity); $\beta$** |  |  |  |  |
| Black | 42,962 | 0.279 (0.177 to 0.381) | $8.35 \times 10^{-8}$ | $8.35 \times 10^{-7}$ |
| South Asian | 42,962 | 0.070 (-0.017 to 0.157) | 0.116 | 0.194 |
| Chinese | 42,962 | 0.131 (-0.018 to 0.280) | 0.0860 | 0.194 |
| Mixed | 42,962 | 0.056 (-0.059 to 0.172) | 0.340 | 0.485 |
| **WMH lesion count; IRR** |  |  |  |  |
| Black | 43,532 | 1.16 (1.10 to 1.22) | $2.70 \times 10^{-8}$ | $2.70 \times 10^{-7}$ |
| South Asian | 43,532 | 1.07 (1.03 to 1.13) | 0.00195 | 0.00651 |
| Chinese | 43,532 | 1.09 (1.01 to 1.18) | 0.0265 | 0.0585 |
| Mixed | 43,532 | 1.02 (0.96 to 1.08) | 0.521 | 0.579 |
| **ePVS count; IRR** |  |  |  |  |
| Black | 43,474 | 1.19 (1.02 to 1.38) | 0.0235 | 0.0570 |
| South Asian | 43,474 | 0.83 (0.72 to 0.95) | 0.00620 | 0.0310 |
| Chinese | 43,474 | 0.84 (0.67 to 1.07) | 0.156 | 0.260 |
| Mixed | 43,474 | 0.92 (0.77 to 1.10) | 0.344 | 0.463 |

**Table 5.** Exploratory focal-lesion comparisons. **Matched sample; negative-binomial models adjusted for age, sex and imaging center**

| Group vs White | Estimate (95% CI) | P | q |
| --- | --- | --- | --- |
| **Lacune count, IRR** |  |  |  |
| South Asian | 1.16 (0.73–1.83) | 0.5319 | 0.5910 |
| Black | 1.31 (0.80–2.14) | 0.2870 | 0.4100 |
| Chinese | 0.46 (0.20–1.06) | 0.0698 | 0.2328 |
| Mixed | 0.81 (0.45–1.45) | 0.4785 | 0.5910 |
| **CMB count, IRR** |  |  |  |
| South Asian | 1.25 (0.75–2.08) | 0.3890 | 0.9644 |
| Black | 1.13 (0.64–2.00) | 0.6656 | 0.9644 |
| Chinese | 0.94 (0.45–1.96) | 0.8741 | 0.9644 |
| Mixed | 0.93 (0.49–1.74) | 0.8093 | 0.9644 |

Income-domain deprivation and vascular measures were examined in adjustment ladders. Sample sizes change across stages. Same-sample comparisons are identified where available; detailed WMH segmentation sensitivity analyses are reported in the supplement, and the full sample ladder remains a changing-sample comparison. Adjustment was not interpreted as a causal explanation or a percentage of an ethnoracial-group difference explained.^20,34^

Lacune and CMB counts were modeled with negative-binomial regression adjusted for age, sex and imaging center; all ten pairwise contrasts are reported in Tables S2 and S3. The 0–2 automated-marker score assigned one point each for log WMH above the full-cohort upper quartile (8.075275) and ePVS count >2. The 0–4 multimarker score added one point each for any CMB and any lacune; missing markers were not scored as absent. Both scores used fixed full-cohort thresholds. These study-specific thresholds differ from the visual-rating cutoffs of the conventional total SVD score.^17^ Proportional-odds models included age, sex and imaging center (Table 6; Figure 2). Score definitions and component frequencies are reported in Table S4, complete matched-sample pairwise estimates in Table S5, and proportional-odds diagnostics in Table S6. The composites are secondary imaging indices, not validated clinical risk scores.

**Figure 2.**
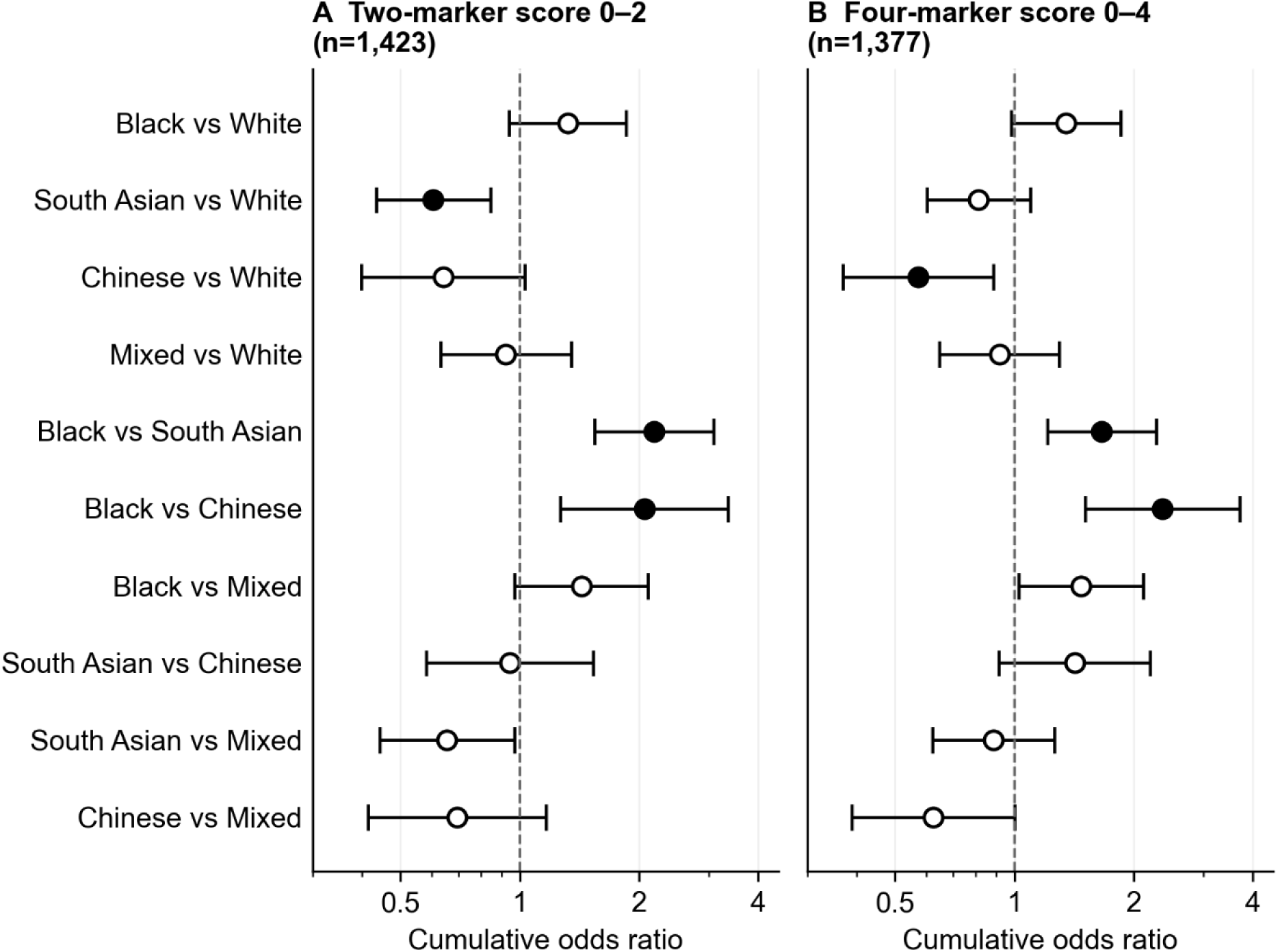
All ten pairwise comparisons for the two-marker and four-marker composite scores in the matched sample. A, 0–2 WMH/ePVS score (model n=1,423). B, 0–4 score additionally including CMB and lacunes (model n=1,377). Proportional-odds models include age, sex and imaging center. Filled points indicate q<0.05 after Benjamini–Hochberg correction across ten pairs within each score; open points indicate q≥0.05. Horizontal lines show 95% CIs on a logarithmic cumulative odds-ratio axis; the dashed line marks an odds ratio of 1. The scores are study-specific imaging indices, not validated clinical risk scores. Contrasts compare the first named group with the second. CMB indicates cerebral microbleeds; ePVS, enlarged perivascular spaces; WMH, white matter hyperintensities.

**Table 6.** Composite cSVD burden in the matched sample. Cumulative odds ratios compare each named group with White participants. Models adjust for age, sex and imaging center.

| Group | OR (95% CI) | P | q |
| --- | --- | --- | --- |
| South Asian | 0.61 (0.43–0.84) | 0.0031 | 0.0117 |
| Black | 1.32 (0.94–1.85) | 0.1077 | 0.1539 |
| Chinese | 0.64 (0.40–1.03) | 0.0662 | 0.1178 |
| Mixed | 0.92 (0.63–1.35) | 0.6791 | 0.7546 |

**Four-marker multimarker score 0–4** Model n=1,377. Omnibus ethnoracial P=0.0011.
| Group | OR (95% CI) | P | q |
| --- | --- | --- | --- |
| South Asian | 0.81 (0.60–1.10) | 0.1759 | 0.2198 |
| Black | 1.35 (0.98–1.86) | 0.0633 | 0.1056 |
| Chinese | 0.57 (0.37–0.88) | 0.0121 | 0.0403 |
| Mixed | 0.92 (0.65–1.30) | 0.6270 | 0.6270 |

LE8–brain and brain–cognition associations were examined with ethnoracial-group interactions, group-specific slopes and available pairwise slope differences. Absence of detected moderation was assessed together with slope-difference confidence intervals, not treated as equivalence. Analyses used R 4.3.2. Model specifications and multiplicity families for the reported comparisons are described in Supplementary Methods S-M6, S-M7 and S-M10.

## Results

### Participants and imaging selection

Table 1 presents participant characteristics in the full and matched samples. Table 2 presents adjusted cardiovascular, cSVD and cognitive profiles in the matched sample. Black participants had a lower age- and sex-adjusted LE8 score than White participants (67.7 [95% CI, 66.3–69.1] versus 73.5 [72.3–74.7]; n=1,423). Adjusted global cognitive test scores were lower in Black (−0.76 [−0.90 to −0.62]) and South Asian (−0.57 [−0.69 to −0.45]) than White participants (0.24 [0.13–0.35]; n=1,043). These group profiles provide clinical context, not evidence that vascular differences explain cognitive performance. Processing-speed and executive-function means were also lower in Black and South Asian participants, whereas Chinese participants had a lower executive-function mean only and Mixed participants showed smaller differences (Table 2). In the matched sample, antihypertensive medication use was 32.4% in Black and 31.4% in South Asian participants, compared with 17.7% in White participants. Imaging participation was socially patterned: the composite-IMD standardized mean differences between imaged and eligible never-imaged participants were −0.34 in Black, −0.39 in South Asian and −0.17 in White participants. These comparisons describe selection into imaging rather than the direction or magnitude of bias in a WMH contrast (Supplementary Results S-R1).

### Composite multimarker cSVD burden

Both composite scores showed overall ethnoracial differences in the matched sample: 0–2 P=0.00010 (n=1,423) and 0–4 P=0.0011 (n=1,377) in age-, sex- and center-adjusted models. For the 0–4 score, Black participants had higher odds of greater burden than South Asian (OR 1.66; 95% CI, 1.21–2.28; q=0.0081) and Chinese participants (OR 2.36; 1.51–3.70; q=0.0018), and Chinese participants had lower odds than White participants (OR 0.57; 0.37–0.88; q=0.0403).

The Black–White contrast was imprecise and not detected (OR 1.35; 0.98–1.86; q=0.1056), as was the Black–Mixed contrast (OR 1.47; 1.03–2.11; q=0.0888) (Table 6; Figure 2; Table S5). The 0–2 score also identified greater burden in Black than South Asian (OR 2.18; 95% CI, 1.54– 3.08; q=0.00010) and Chinese participants (OR 2.06; 1.27–3.35; q=0.0117). South Asian participants had lower odds than White participants (OR 0.61; 0.43–0.84; q=0.0117). The Black– White contrast was not detected for this score (OR 1.32; 0.94–1.85; q=0.1539). The full-sample 0–2 omnibus test was P=3.4×10⁻⁷ (n=44,519). The scores were correlated in matched participants with both scores available (Spearman ρ=0.817; n=1,378), but their group contrasts differed. Complete-case samples vary, so this is not a formal test of the additional contribution of focal lesions (Table S5).

### Individual markers: higher WMH volume in Black participants

In the full sample, the main UBO analysis showed higher WMH volume in Black than White participants (β=0.209; 95% CI, 0.110–0.308; P=3.5×10⁻⁵; q=0.0002; n=44,002). Black participants also had higher UBO WMH volume than South Asian, Chinese and Mixed participants in the all-pairwise analysis (Figure 1; Table S7). After income-domain IMD adjustment, the Black–White estimate was β=0.178 (95% CI, 0.077–0.279; P=0.00055; q=0.0028; n=42,962), and all four Black-versus-group UBO contrasts remained below the within-marker FDR threshold (Table S7).

In sensitivity analyses, the standard UK Biobank BIANCA measure gave the same principal Black–White full-sample contrast (β=0.311; 95% CI, 0.211–0.412; P=1.1×10⁻⁹; q=1.1×10⁻⁸; n=44,002), including after income-domain IMD adjustment (β=0.279; 95% CI, 0.177–0.381; P=8.4×10⁻⁸; q=8.4×10⁻⁷; n=42,962). Secondary group contrasts were not uniformly stable across segmentation methods and are detailed in the supplement (Figure S2).

In the matched sample (N=1,426), the main UBO Black–White estimate was β=0.108 (95% CI, −0.018 to 0.234; P=0.0938; q=0.1735; n=1,411; Table S1). The interval included no difference. Because the analysis levels overlap and differ principally in their White comparison group, this result is interpreted as sensitivity to cohort implementation and precision rather than an independent replication test.

In the matched-sample BIANCA sensitivity analysis, the same-sample base estimate was 0.248 (95% CI, 0.103–0.393) and the fully adjusted estimate was 0.199 (0.051–0.346; source-ladder q=0.0849) (n=1,083 in both models). This reduction after additional adjustment did not retain multiplicity-controlled significance. The full-sample adjustment ladder uses changing samples and is not a common-sample decomposition of Table 4 (Table S8; Figure S3).

### Marker-specific findings beyond total WMH volume

South Asian participants had fewer ePVS than White participants in the full sample (IRR 0.82; 95% CI, 0.72–0.93; P=0.0026; q=0.0128; n=44,519). The corresponding matched-sample estimate was IRR 0.87 (95% CI, 0.72–1.06; P=0.166; q=0.3317; n=1,423) and was less precise (Table 4 and Table S1). Black participants had more ePVS than South Asian participants at both levels (full sample IRR 1.39; 95% CI, 1.14–1.69; q=0.0099; n=44,519; matched IRR 1.41; 1.15– 1.73; q=0.011; n=1,423). The matched Black–White contrast was not detected in the main model (IRR 1.22; 95% CI, 0.99–1.51; q=0.1495) but was after income-domain IMD adjustment (IRR 1.38; 1.11–1.71; q=0.0203; n=1,372) (Table S7). These findings do not imply lower overall cSVD burden in any group.

WMH lesion counts conditional on WMH volume were higher in South Asian participants at both levels: IRR 1.08 (95% CI, 1.03–1.13; P=0.0012; q=0.0045; n=44,580) in the full sample and IRR 1.13 (95% CI, 1.06–1.21; P=0.00038; q=0.0019; n=1,425) in the matched sample. Black participants also had higher conditional lesion counts: IRR 1.18 (95% CI, 1.12–1.24; P=8.2×10⁻¹⁰; q=8.2×10⁻⁹; n=44,580) and IRR 1.20 (95% CI, 1.12–1.29; P=5.2×10⁻⁷; q=5.2×10⁻⁶; n=1,425), respectively. Count differences describe the distribution of lesions at a given measured volume; they do not demonstrate differences in tissue injury mechanisms.

In the matched sample, South Asian participants had lower main-analysis UBO WMH volume than White participants (β=−0.163; 95% CI, −0.282 to −0.044; P=0.0073; q=0.0245; n=1,411), including after income-domain IMD adjustment (β=−0.168; 95% CI, −0.289 to −0.046; P=0.0071; q=0.0258; n=1,361). This matched-sample contrast was not supported by the BIANCA sensitivity analysis, and the full-sample UBO South Asian–White contrast was not detected (Table 4; Table S7; Figure S2).

### Focal lesions in the matched sample

Lacune and CMB comparisons complete the conventional four-marker profile but were sparse. In the matched-sample lacune analysis, no overall ethnoracial difference was detected (omnibus P=0.075; n=1,425). The Black–White count estimate was IRR 1.31 (95% CI, 0.80–2.14; P=0.287; q=0.410). The largest contrast, Black versus Chinese (IRR 2.83; 1.22–6.57; P=0.015), did not survive correction (q=0.146); no lacune contrast among the ten group pairs did (Table 5; Table S2).

CMB were present in 137/1,380 participants with SWI, with 136/1,379 in the model after the sparse-center exclusion. No ethnoracial-group difference in CMB count was detected (omnibus P=0.84); the Black–White IRR was 1.13 (95% CI, 0.64–2.00; P=0.666; q=0.964; n=1,379), and no pairwise CMB contrast passed correction (Table 5; Table S3). The absence of a detected contrast does not establish equal CMB burden, particularly with sparse events. Threshold analyses are supplementary (Table S9).

### Cardiovascular-health associations, cognition and uncertainty in moderation

Better cardiovascular health was associated with lower matched-sample UBO WMH volume (β=−0.129 per SD higher LE8; 95% CI, −0.171 to −0.087; q=9.4×10⁻⁸; n=1,408) and fewer WMH lesions (IRR 0.89; 0.86–0.93; q=3.6×10⁻⁷; n=1,422; Table S10). These pooled associations provide vascular context; they do not show that LE8 accounts for the ethnoracial contrasts.

No moderation of the principal LE8 total–brain or brain–cognition associations by ethnic group was detected after the family-specific corrections. Confidence intervals nevertheless included appreciable slope differences (Table S10). For LE8 total and main UBO WMH, the Black-minus-White slope difference was 0.073 (95% CI, −0.044 to 0.190; source-family q=0.9978; n=1,408). For UBO WMH and global cognition, the Black-minus-White difference was −0.034 (95% CI, −0.205 to 0.138; source-family q=0.9963) and the South Asian-minus-White difference was −0.175 (−0.350 to −0.000; source-family q=0.9963) (n=1,035). These intervals do not establish common slopes.

Regional estimates are reported in Table S11. In the matched sample, South Asian participants had lower periventricular WMH volume than White participants (β=−0.203; SE 0.061; P=0.00084; q=0.0033; n=1,411), whereas deep WMH contrasts were not detected; basal ganglia ePVS counts were nominally higher in Black participants (IRR 1.29; 95% CI, 1.03–1.61; P=0.024) but did not pass correction (q=0.080), and no centrum semiovale contrast was detected. Cognitive test-performance differences across adjustment stages are reported in Table S12.

Neither PSMD nor DDF showed a detected omnibus ethnoracial-group difference in the matched sample (PSMD P=0.29; DDF P=0.17; n=1,364); in the full sample, PSMD was higher in South Asian than White participants (β=1.35×10⁻⁵; 95% CI, 4.3×10⁻⁶ to 2.27×10⁻⁵; q=0.040; n=42,967) and DDF was lower in Black participants (β=−0.0036; −0.0060 to −0.0012; q=0.023; n=42,969) (Table S13). These secondary measures do not establish equivalent or different white matter integrity across groups.

Better cardiovascular health was also associated with lower aggregate cSVD burden. In the full sample, each SD higher LE8 total score was associated with lower odds of greater two-marker burden (cumulative OR 0.89; 95% CI, 0.87–0.91; q=1.8×10⁻³⁰; n=44,474), with the blood-pressure component showing the strongest association (OR 0.82 per SD; 0.80–0.84; q=6.9×10⁻⁷⁴; n=43,787). In the matched sample, the LE8 total association with the four-marker score was not detected (OR 0.95; 95% CI, 0.85–1.07; q=0.77; n=1,374), whereas antihypertensive medication use, a marker of treated vascular disease, was associated with greater four-marker burden (OR 1.66; 1.29–2.14; q=0.0021; n=1,375) (Table S14).

## Discussion

This UK Biobank study identified ethnoracial variation in both aggregate cSVD burden and individual MRI manifestations. Both composite scores showed overall group differences and consistently distinguished Black from South Asian and Chinese participants, whereas Black– White composite contrasts were imprecise. The constituent markers did not form a uniform hierarchy: the clearest Black–White difference was higher WMH volume in the full sample, retained after measured deprivation adjustment and with an alternative segmentation; ePVS showed a different pattern, with fewer ePVS in South Asian than White participants and more in Black than South Asian participants; and focal-lesion comparisons were inconclusive. Better LE8 cardiovascular health was associated with lower WMH burden across groups without detected moderation and with lower aggregate burden in the full sample, and Black participants had the least favorable LE8 scores. The main implication is that conclusions about ethnoracial differences in cSVD depend partly on how vascular brain injury is represented: a composite summarizes co-occurring burden, whereas component analyses reveal marker-specific patterns that aggregation can obscure.^1,3,11^

The Black–White WMH finding is consistent with evidence from US cohorts and extends the comparison to a UK imaging setting.^8–10^ Less favorable measured cardiovascular health in Black participants in the matched sample coexisted with greater full-sample WMH burden (Tables 1, 2 and 4). Smaller adjusted estimates are compatible with contributions from vascular and social exposures, but do not identify a causal explanation. Area deprivation and current vascular measurements incompletely represent accumulated exposure, healthcare access, environmental conditions, structural disadvantage and discrimination.^19,35,36^ The remaining contrast is unexplained by measured factors, not evidence of an intrinsic ethnic susceptibility.

Selection into imaging was more strongly socially patterned in Black and South Asian participants. The selection analysis documents an important limitation of generalizing absolute burden and group contrasts beyond the imaging participants; it does not provide bias-corrected estimates.^37,38^ The overlapping matched sample changes the White comparison group and affords focal-lesion assessment, rather than independent replication. Its imprecise Black–White UBO result therefore qualifies the full-sample finding. Conditioning on participation can also distort associations in either direction.^39^

The component measures are essential to interpreting what the composite and WMH results do, and do not, represent. Fewer ePVS in South Asian participants did not imply lower overall injury, particularly alongside higher WMH lesion counts at a given volume. SABRE similarly illustrates that WMH distributions and risk-factor associations need not follow one pattern.^14^ Lacunes and CMB capture focal ischemic and hemorrhagic manifestations not described by WMH volume alone.^4,16^ Their blinded, reviewed assessment, including radiologist validation, is detailed in Table 3. Inconclusive focal-lesion comparisons do not make these measurements uninformative, but they neither establish equal burden nor demonstrate improved prediction.

Regional WMH and ePVS analyses describe anatomical distribution, and PSMD and DDF are complementary microstructural measures with no detected omnibus group difference in the matched sample.

The two composites answer complementary questions. The 0–2 score characterizes the co-occurrence of elevated automated WMH and ePVS measures across the large imaging sample; the 0–4 score additionally captures focal ischemic and hemorrhagic lesions in the matched sample. Both showed higher scores in Black than South Asian and Chinese participants, but the South Asian–White and Chinese–White patterns differed between scores and the Black–White contrasts were imprecise. Composite and component measures are therefore complementary rather than competing. The correlation between the two scores does not establish interchangeability or incremental prognostic value, and their upper-quartile automated thresholds make them study-specific indices distinct from the conventional visual-rating total SVD score.^17,40^

The association between better LE8 cardiovascular health and lower WMH burden provides a clinically relevant exposure context for the imaging differences, and the same direction was seen for aggregate burden on the full-sample two-marker score, where the blood-pressure component carried the strongest association.^31^ Antihypertensive medication use was associated with greater four-marker burden in the matched sample; this marks treated vascular disease rather than an effect of treatment. These results support prevention research across groups without attributing a group contrast to a single current risk measure. No ethnoracial moderation of the principal LE8– brain or brain–cognition associations was detected after correction, but the intervals do not establish equivalent associations.^14,41–43^ These are cross-sectional associations; neither they nor the group cognitive profiles establish mediation or clinical prognosis. The cognitive means in Table 2 did not follow the WMH gradient: South Asian participants had lower matched-sample WMH volume yet cognitive deficits similar to those in Black participants, the pooled WMH– cognition coefficient was null (Table S10), and the deficits were unchanged by education adjustment and only partly attenuated in Black participants by broader socioeconomic adjustment (Table S12). The executive-function deficit was large in South Asian, Black and

Chinese participants, whereas Chinese memory and processing-speed means were not lower, a pattern more consistent with test-format and language effects than with vascular injury.^32,33^ Black participants nevertheless combined higher WMH volume, less favorable LE8 scores and lower cognitive means, so downstream cognitive consequences of cSVD in this group warrant prospective study, although their cross-sectional WMH–cognition slope was close to zero (Table S10).

The study combines a large imaging sample with focused multimarker assessment in an ethnoracially diverse matched sample, including Chinese and Mixed participants often absent from named-group comparisons. Total and regional WMH, lesion counts, total and regional ePVS, PSMD and DDF complement lacune and CMB assessments. Both composite definitions, all ten group pairs, formal interaction tests and explicit model denominators are retained.

Socioeconomic and vascular adjustment and the imaging-selection analysis place the findings in a broader exposure context. Sensitivity analyses with an alternative WMH segmentation supported the principal full-sample contrast while identifying method-dependent secondary findings; neither method is a reference standard.^44^

The cross-sectional design cannot establish progression, mediation or incident stroke risk. The large full sample does not remove uncertainty from the smaller South Asian, Black, Chinese and Mixed samples, sparse focal-lesion events or changing complete-case samples. Residual social confounding, possible measurement non-equivalence and proportional-odds diagnostics also limit inference. Broad self-reported identities conceal substantial within-group heterogeneity.^45,46^ Neither group membership nor a single MRI measure should substitute for individual clinical assessment.

Ethnoracial differences in this study were evident in overall multimarker cSVD burden, but their pattern depended on the constituent MRI manifestation. For cerebrovascular research, the practical implication is to report an aggregate score alongside its components and the measured vascular and social context. The consistent LE8–WMH association across groups identifies cardiovascular health as a tractable target for prevention research in diverse populations, although the cross-sectional design cannot show that improving LE8 reduces cSVD. Longitudinal studies should test whether these profiles predict incident stroke or cognitive decline and whether addressing modifiable exposures reduces injury. Diverse VCID and other prospective multimarker studies provide a setting for that next step.^47,48^

## Data Availability

This research was conducted using the UK Biobank Resource under application 98013. Individual participant data from UK Biobank cannot be redistributed directly by the authors under the terms of UK Biobank access. Bona fide researchers may apply for access to the same data through UK Biobank. Derived analysis code and nonidentifiable aggregate summary outputs may be made available on reasonable request to the corresponding author.

## Non-standard Abbreviations and Acronyms

BIANCA: Brain Intensity AbNormality Classification Algorithm
CMB: cerebral microbleeds
cSVD: cerebral small vessel disease
DDF: difference in distribution functions
EMM: estimated marginal mean
ePVS: enlarged perivascular spaces
FDR: false discovery rate
ICV: intracranial volume
IMD: Index of Multiple Deprivation
IRR: incidence rate ratio; here, a ratio of expected lesion counts
LE8: Life’s Essential 8
PSMD: peak width of skeletonized mean diffusivity
STRIVE: Standards for Reporting Vascular Changes on Neuroimaging
SWI: susceptibility-weighted imaging
UBO Detector: automated WMH segmentation pipeline
WMH: white matter hyperintensities

## Acknowledgments

We thank the UK Biobank participants and staff. This research was conducted using the UK Biobank Resource under application 98013.

No non-author contributors are named. During manuscript preparation, the authors used ChatGPT (OpenAI) and Claude (Anthropic) for editing and formatting. The authors verified all content and are responsible for all analyses and all manuscript content.

Nikita K. Husein had full access to all of the data in the study and takes responsibility for the integrity of the data and the accuracy of the data analysis.

## Author contributions

concept and design, Husein, Wen, P. S. Sachdev; acquisition, analysis, or interpretation of data, Husein, Ho, Lin, Chan, Wen; drafting of the manuscript, Husein; critical review for important intellectual content, all authors; statistical analysis, Husein, Chan; obtained funding, P. S. Sachdev, Wen; administrative, technical, or material support, Ho, Lin, Hayashi, S. P. Sachdev, Hassanzadeh, Jiang; supervision, Wen, P. S. Sachdev.

## Supplemental Material

Supplementary Methods S-M1–S-M10; Supplementary Results S-R1–S-R6; Figures S1–S3; Tables S1–S14; STROBE and Disparities Research Guidelines checklists (also supplied as separate files).

